# Using species richness calculations to model the global profile of unsampled pathogenic variants: Examples from *BRCA1* and *BRCA2*

**DOI:** 10.1101/2022.11.09.22282146

**Authors:** Nandana D. Rao, Brian H. Shirts

## Abstract

There have been many surveys of genetic variation in *BRCA1* and *BRCA2* to identify variant prevalence and catalogue population specific variants, yet none have evaluated the magnitude of unobserved variation. We applied species richness estimation methods from ecology to estimate “variant richness” and determine how many germline pathogenic *BRCA1/2* variants have yet to be identified and the frequency of these missing variants in different populations. We also estimated the prevalence of germline pathogenic *BRCA1/2* variants and identified those expected to be most common. Data was obtained from a literature search including studies conducted globally that tested the entirety of *BRCA1/2* for pathogenic variation. Across countries, 45% to 88% of variants were estimated to be missing, i.e., present in the population but not observed in study data. Estimated variant frequencies in each country showed a higher proportion of rare variants compared to recurrent variants. The median prevalence estimate of *BRCA1/2* pathogenic variant carriers was 0.64%. *BRCA1* c.68_69del is likely the most recurrent *BRCA1/2* variant globally due to its estimated prevalence in India. Modeling variant richness using ecology methods may assist in evaluating clinical targeted assays by providing a picture of what is observed with estimates of what is still unknown.

## Introduction

*BRCA1* and *BRCA2* are among the most studied human genes, with many thousands of variants in each gene defined and dozens of populations surveyed. Although studies report on pathogenic variants observed, none have evaluated what is potentially missing in these surveys. Understanding pathogenic variation in these genes is important because these variants increase risk for several cancers including breast, ovarian, prostate, and pancreatic [1]. Knowledge about carrier status has useful clinical implications, as individuals who learn about their pathogenic variation early through genetic testing can engage in more frequent screening to increase the odds of early cancer detection and undergo prophylactic surgery to decrease future cancer risk.

Many recent studies have estimated the overall prevalence of pathogenic *BRCA1/2* variant carriers in specific populations. Population genetic screening studies in the United States have reported prevalence estimates ranging from 0.5% to 0.7% [2,3,4]. Higher prevalence has been seen among Ashkenazi Jewish populations, reaching 2% [2]. These estimates provide insight into the burden of *BRCA1/2*-related hereditary cancer and indicate that the number of individuals who might benefit from genetic testing and subsequent preventive measures is substantial. However, less is known about the prevalence of *BRCA1/2* pathogenic variant carriers in other populations around the world.

Understanding the types of pathogenic *BRCA1/2* variation in a given population can also be useful for informing genetic screening strategies. For instance, if certain pathogenic variants are found more commonly in a particular population, this can lead to the development of population-specific screening strategies [5,6]. Alternatively, if there appears to be a wide range of variation, more comprehensive sequencing may better detect pathogenic variant carriers. There have been extensive efforts of large consortia to understand the global spectrum of pathogenic *BRCA1/2* variants, including the most recurrent variants in different regions around the globe [7]. However, many statistical analyses of these results are limited by inconsistencies in ascertainment strategies, variant testing methods, and an inability to account for variation that has not yet been observed.

The purpose of this study was to model the prevalence of germline pathogenic *BRCA1/2* variants in different populations around the world and predict their allele frequency spectrum. Specifically, we sought to determine how many germline pathogenic *BRCA1/2* variants have yet to be identified in well-studied countries and how common unreported variants are in their respective populations. We also compiled a list of the most recurrent observed variants in several countries. Given the clinical significance of *BRCA1/2*, it is important to understand the number of people who may benefit from genetic testing and what spectrum of pathogenic variants should be expected in different places around the world.

## Materials and Methods

### Data Sources

PubMed was used to search for studies published between January 1999 and March 2020 that involved screening individuals for pathogenic variants in *BRCA1* and *BRCA2*. Search terms included “*BRCA1*”, “*BRCA2*”, “breast cancer”, “ovarian cancer”, “population screening”, “gene sequencing”, and “direct sequencing”. The resulting studies and their references were examined and only those that tested the entirety of these genes for pathogenic variation were included in later analyses. Studies that targeted specific variants or that used methods capable of detecting only a subset of variants were excluded. For our analyses, variants were classified as pathogenic using classifications from individual studies based on ACMG PVS1 criteria for dominant hereditary breast and ovarian cancer risk in the *BRCA1* and *BRCA2* genes or likely pathogenic or pathogenic classifications in ClinVar [8]. The following accession version numbers were used for *BRCA1* and *BRCA2*, respectively: NM_007294.3 and NM_000059.3. Detailed search and selection procedures are shown in S1 Fig.

### Unique Variant Estimation Methods

We sought to apply species richness estimation methods from field ecology to estimate the number of unique pathogenic variants in a given gene that are present in a population and the relative frequency of these variants. Briefly, species methods look at the number of unique species and their frequencies in a <u>sample</u> of the environment to estimate the <u>total</u> number of unique species in the same environment. “Species” can be defined broadly (e.g., words in a book, bugs in software programs, alleles in genetic code), so these methods have many applications [9]. Underlying assumptions of species richness estimate methods are [10,11,12]:

1. Individual representatives of a species are independently and randomly sampled from a population.
2. Species are distributed uniformly in a specific catchment area.
3. Species distributions can be mathematically defined.

For all practical purposes, similar assumptions apply for estimating the total number of unique variants from a population sample of variants: 1) Most variant assessment studies are blind to variant status before sequencing. To further meet this assumption in our analysis, if multiple individuals from the same family were sequenced, only one individual was selected for inclusion. Additionally, data was included from studies performing sequencing rather than targeted testing to meet the criteria of random ascertainment. 2) Population substructure is always present in human populations and ecology. Since human populations are relatively large and variant status is unknown before sequencing, variant estimates should perform as well as species estimates with regards to this assumption. For our purposes, we used country as a catchment area given its public health relevance and because most study samples focused on a particular country. 3) We illustrate below that variant distributions follow patterns similar to species distributions.

There are several specific methods for estimating species richness, each with different strengths and different parameters for modeling distributions [13]. The Chao 1984 method [14] is a relatively simple and straightforward method that has been shown to give an accurate lower bound estimate. It assumes that variants (or species) that occur rarely provide the most information about the number of missing variants (or species). Importantly, this method only uses singletons and doubletons for estimation, so it breaks down when no doubletons occur [15]. We used the Chao method to estimate a lower bound of the total number of unique variants.

There are also several maximum likelihood methods for predicting species richness. Literature on maximum likelihood methods has shown that these give more accurate estimates but can be sensitive to input parameters. The penalized nonparametric maximum likelihood method (pnpml) assumes the number of unique variants fits a mixed Poisson distribution [16]. We chose the pnpml method to estimate the total number of unique variants because it also models the distribution of variant frequencies with estimated mixed Poisson parameters.

### Application of variant estimation to BRCA1/2

Data for *BRCA1/2* variant estimation were extracted from studies that identified at least 40 unique pathogenic variants because estimation techniques were sensitive to sample size and provided more stable estimates with a larger sample [11]. For some countries, data from several studies with unique participants were combined to obtain a larger sample of pathogenic variants. When variant nomenclature differed between studies, common variant names were identified via ClinVar [8] and HGMD [17] so that the frequency of variants could be more accurately determined. The variant data used for each country are listed in S1 Table. Both the Chao 1984 and pnpml method were implemented in R version 3.6.1 using functions from the SPECIES package [18]. Estimation parameters were not adjusted for variant sample size. In addition, as a supplemental analysis, we applied Chao and pnpml methods to *BRCA1/2* gnomAD v2.1 data [19] for populations where at least 40 unique likely pathogenic or pathogenic variants, as identified by ClinVar [8], were observed in these genes.

### Prevalence Calculations

Overall frequency of variation is relevant to estimates of the total number of variants present. Studies from the PubMed search that had at least 100 participants and included both likely hereditary and sporadic cancer cases were used to assess the prevalence of pathogenic *BRCA1/2* variant carriers in each study location. Cases were considered likely hereditary for a variety of reasons such as cancer diagnosis before age 35, bilateral breast cancer before age 50, and/or first-degree relative(s) with breast or ovarian cancer, though classification varied from study to study based on the information provided and, in some instances, cases were excluded from further analysis when the appropriate information was not provided.

For each study meeting the above criteria, the following data was extracted separately for likely hereditary and sporadic cancer cases if available: the number of individuals recruited, the number of individuals tested for pathogenic *BRCA1/2* variation, and the number of pathogenic *BRCA1/2* variant carriers. Extracted datasets with complete information can be found in S2 Table. These data were used to estimate the prevalence of pathogenic *BRCA1/2* variant carriers in each country by maximizing the Horvitz-Thompson pseudo likelihood function described in Whittemore et. al., which takes into account the number of individuals identified with a pathogenic variant among hereditary and sporadic cancer cases separately [20]. For estimation involving breast cancer cases, 0.57 was used as the probability of disease by age 70 given a *BRCA1/2* pathogenic variant [21], while 0.07 was the probability of disease for noncarriers [22]. For ovarian cancer cases, 0.4 and 0.006 were used for these respective probabilities [21,22]. Confidence intervals for all prevalence estimates were calculated using log transformed data.

### Variant Scaling

All pathogenic variants, even founder variants in *BRCA1/2*, were rare in all populations studied. However, for our analysis, “rare” is a relative term used to refer to variants that represent fewer than 5% of the *BRCA1/2* variants in a given population or a given study. Recurrent pathogenic variants were identified using the studies included in the variant estimation analyses and were defined as variants that made up at least 5% of the observed variants for a respective country. The number of carriers of each recurrent variant in each country was estimated using the 2019 population size of the country, as listed by The World Bank [23]. The prevalence estimate used, 0.62%, was determined by averaging the prevalence estimates of *BRCA1/2* pathogenic variant carriers from 3 population screening studies: The Healthy Nevada Project [3], the Geisinger MyCode Community Health Initiative [4], and the Bio*Me* BioBank [2].

## Results

We found that the Chao and pnpml species richness estimation methods could be applied to estimate “variant richness” and the number of missing variants in *BRCA1/2*. The pnpml method was more informative, generating an expected distribution, whereas the Chao method provided only a discrete number of expected variants. A total of 53 studies were included in our analyses (Table 1), with 48 studies providing data for variant count estimation and 11 for *BRCA1/2* prevalence estimation (6 had data used for both estimates). China, Spain, and the United States had the most studies identified meeting inclusion criteria. Of the 15 countries represented in our analyses, 2 were from North America, 1 from South America, 5 from Europe, 6 from Asia, and 1 from Australia.

**Table 1:**
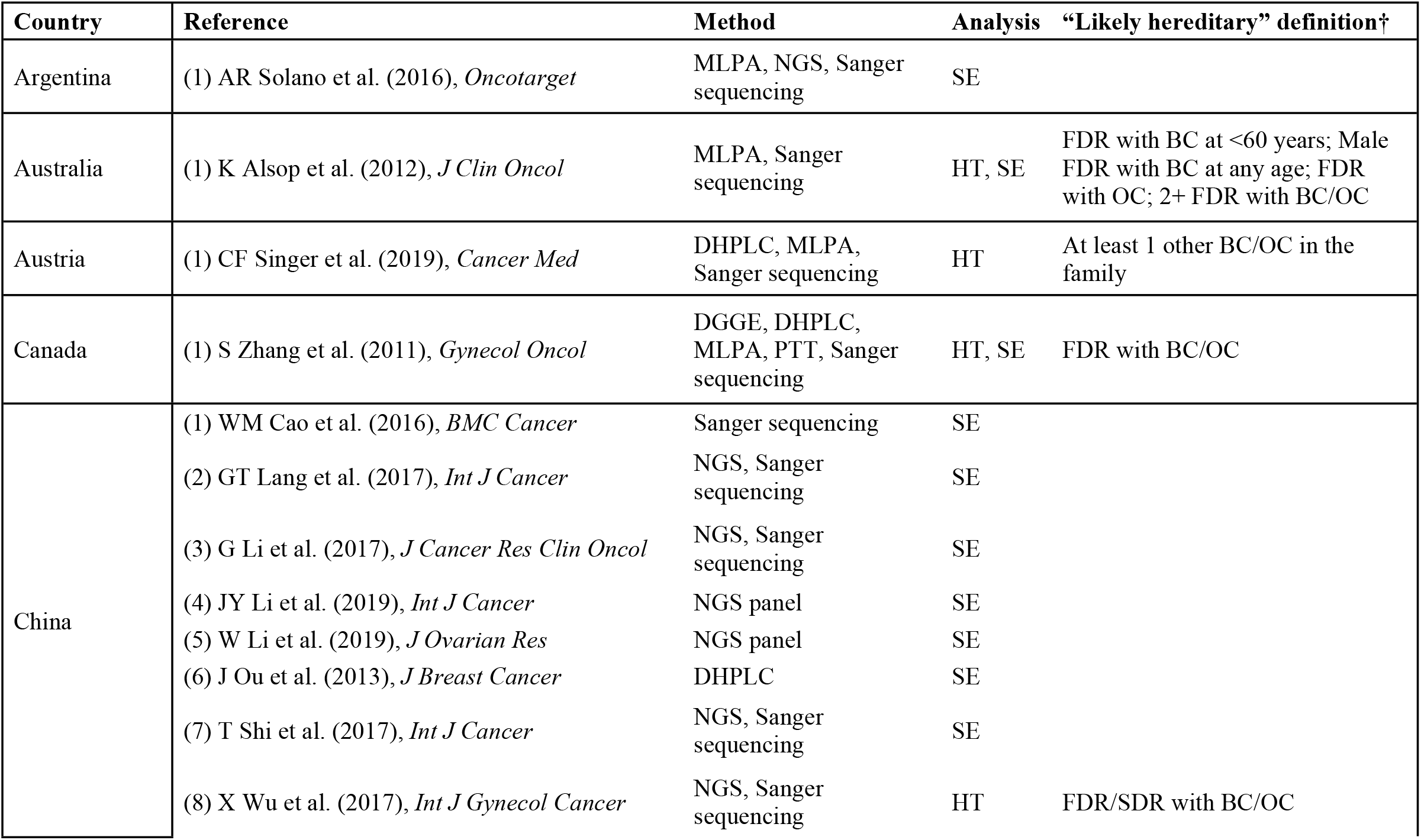
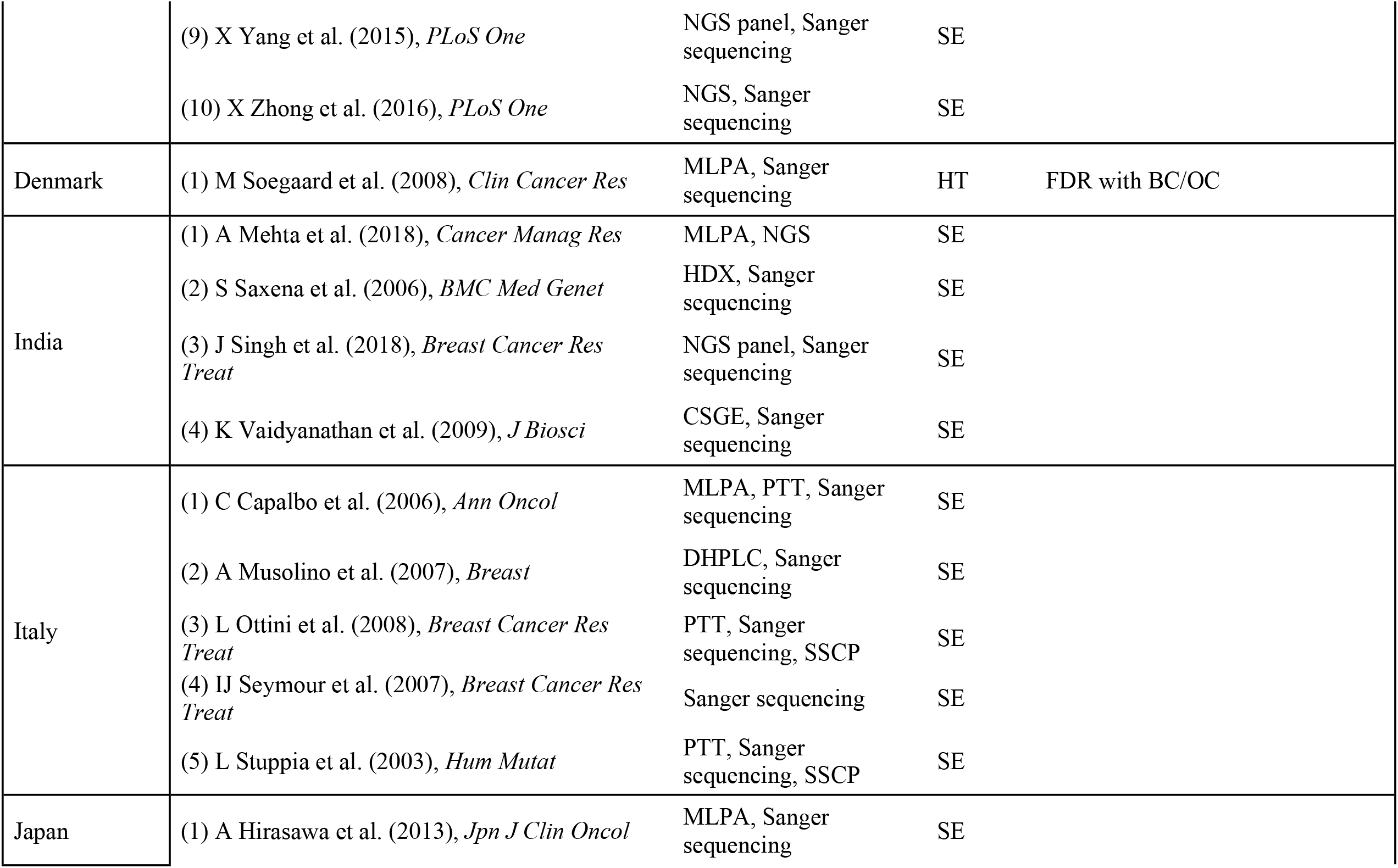
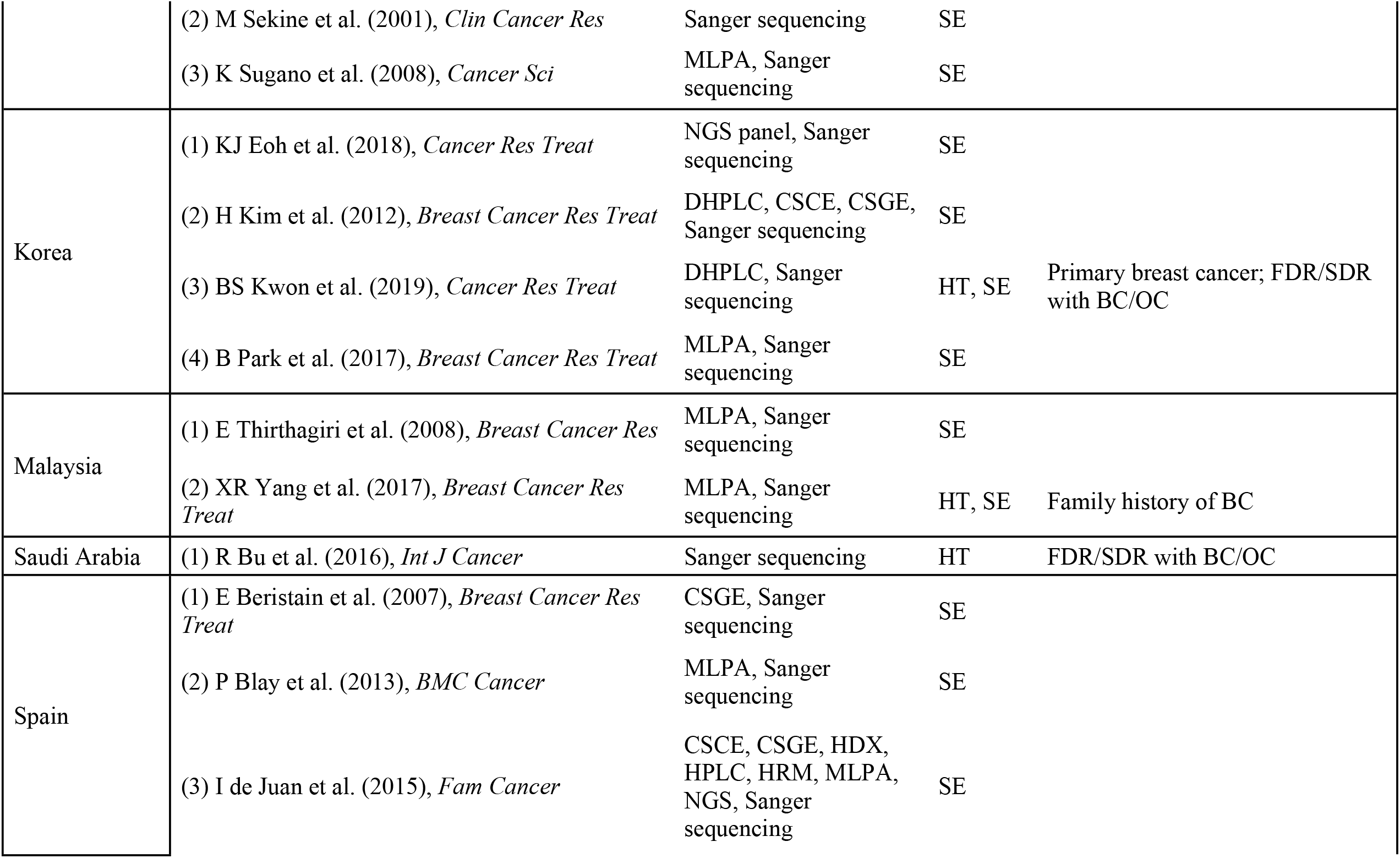
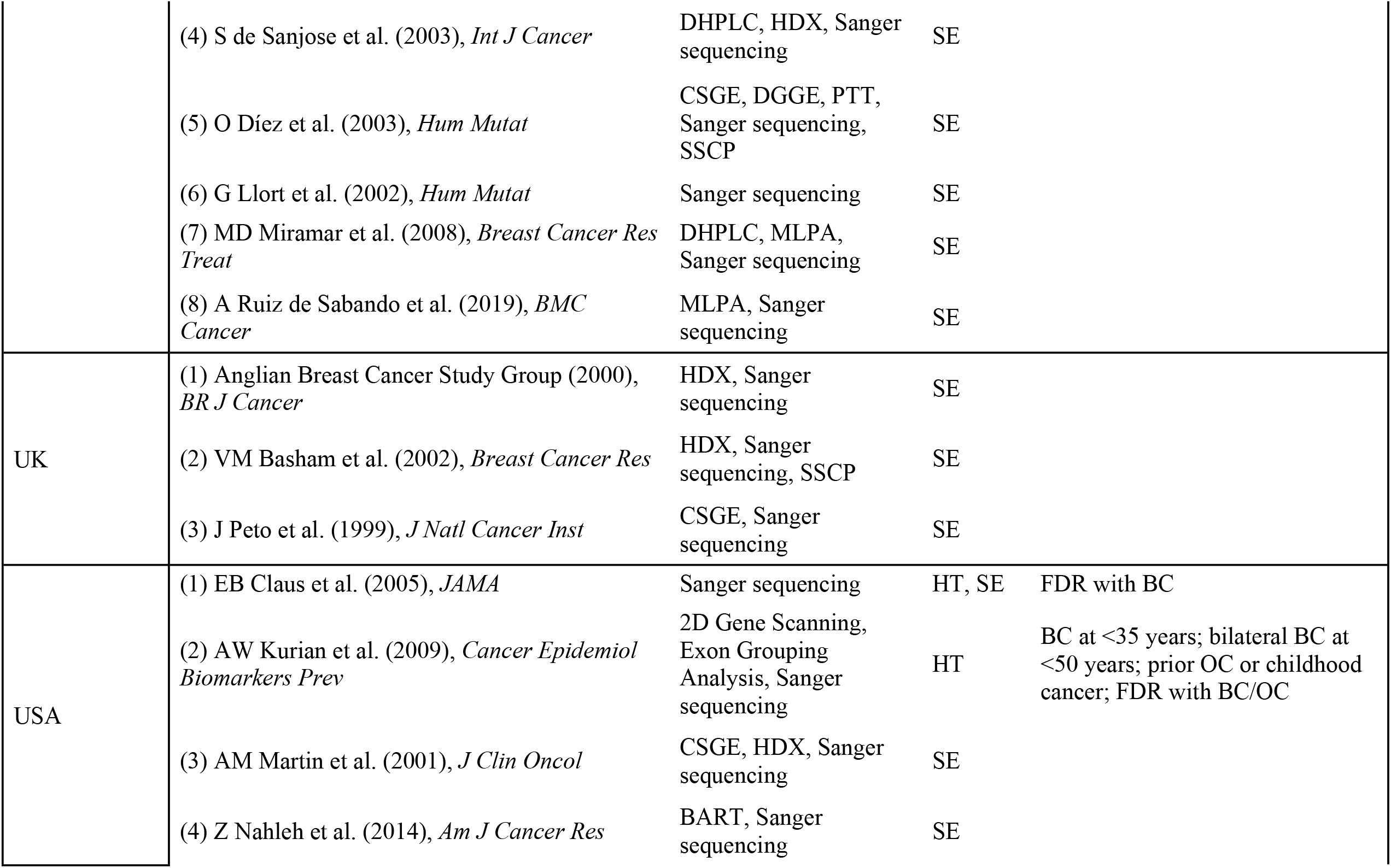
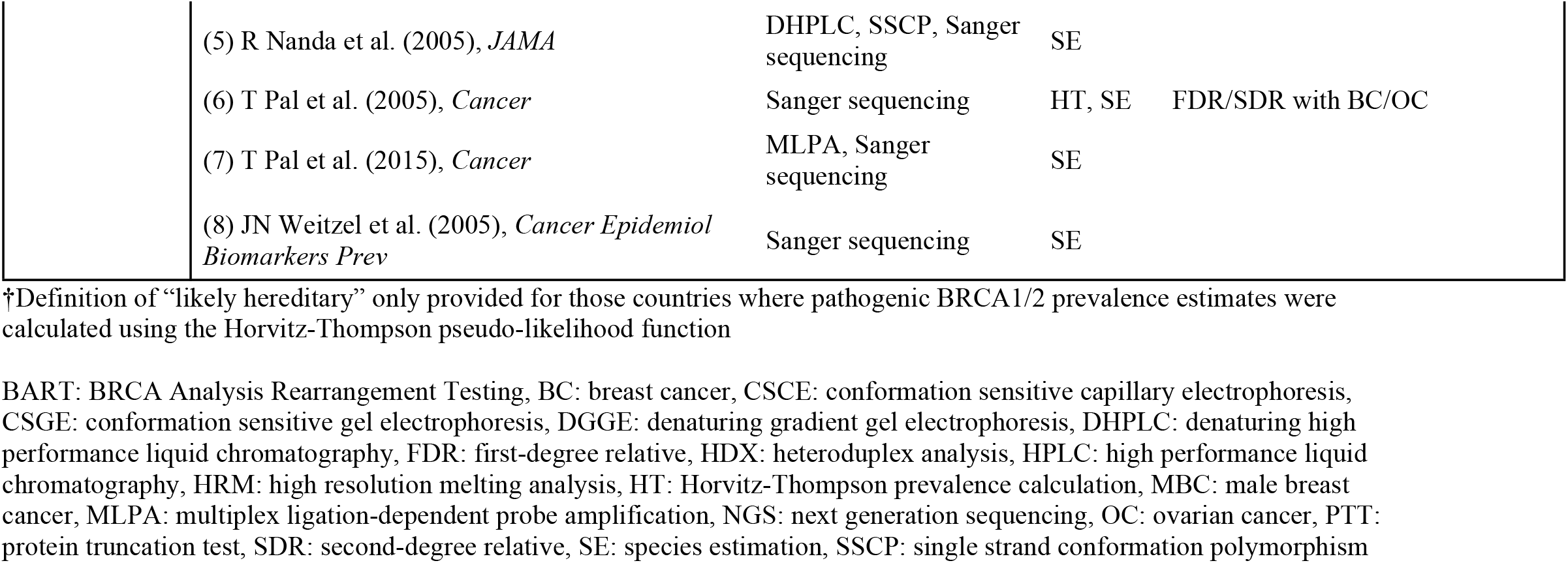
Studies included in analyses.

| Country | Reference | Method | Analysis | “Likely hereditary” definition† |
| --- | --- | --- | --- | --- |
| Argentina | (1) AR Solano et al. (2016), *Oncotarget* | MLPA, NGS, Sanger sequencing | SE |  |
| Australia | (1) K Alsop et al. (2012), *J Clin Oncol* | MLPA, Sanger sequencing | HT, SE | FDR with BC at <60 years; Male FDR with BC at any age; FDR with OC; 2+ FDR with BC/OC |
| Austria | (1) CF Singer et al. (2019), *Cancer Med* | DHPLC, MLPA, Sanger sequencing | HT | At least 1 other BC/OC in the family |
| Canada | (1) S Zhang et al. (2011), *Gynecol Oncol* | DGGE, DHPLC, MLPA, PTT, Sanger sequencing | HT, SE | FDR with BC/OC |
| China | (1) WM Cao et al. (2016), *BMC Cancer* | Sanger sequencing | SE |  |
|  | (2) GT Lang et al. (2017), *Int J Cancer* | NGS, Sanger sequencing | SE |  |
|  | (3) G Li et al. (2017), *J Cancer Res Clin Oncol* | NGS, Sanger sequencing | SE |  |
|  | (4) JY Li et al. (2019), *Int J Cancer* | NGS panel | SE |  |
|  | (5) W Li et al. (2019), *J Ovarian Res* | NGS panel | SE |  |
|  | (6) J Ou et al. (2013), *J Breast Cancer* | DHPLC | SE |  |
|  | (7) T Shi et al. (2017), *Int J Cancer* | NGS, Sanger sequencing | SE |  |
|  | (8) X Wu et al. (2017), *Int J Gynecol Cancer* | NGS, Sanger sequencing | HT | FDR/SDR with BC/OC |
|  | (9) X Yang et al. (2015), *PLoS One* | NGS panel, Sanger sequencing | SE |  |
|  | (10) X Zhong et al. (2016), *PLoS One* | NGS, Sanger sequencing | SE |  |
| Denmark | (1) M Soegaard et al. (2008), *Clin Cancer Res* | MLPA, Sanger sequencing | HT | FDR with BC/OC |
| India | (1) A Mehta et al. (2018), *Cancer Manag Res* | MLPA, NGS | SE |  |
|  | (2) S Saxena et al. (2006), *BMC Med Genet* | HDX, Sanger sequencing | SE |  |
|  | (3) J Singh et al. (2018), *Breast Cancer Res Treat* | NGS panel, Sanger sequencing | SE |  |
|  | (4) K Vaidyanathan et al. (2009), *J Biosci* | CSGE, Sanger sequencing | SE |  |
| Italy | (1) C Capalbo et al. (2006), *Ann Oncol* | MLPA, PTT, Sanger sequencing | SE |  |
|  | (2) A Musolino et al. (2007), *Breast* | DHPLC, Sanger sequencing | SE |  |
|  | (3) L Ottini et al. (2008), *Breast Cancer Res Treat* | PTT, Sanger sequencing, SSCP | SE |  |
|  | (4) IJ Seymour et al. (2007), *Breast Cancer Res Treat* | Sanger sequencing | SE |  |
|  | (5) L Stuppia et al. (2003), *Hum Mutat* | PTT, Sanger sequencing, SSCP | SE |  |
| Japan | (1) A Hirasawa et al. (2013), *Jpn J Clin Oncol* | MLPA, Sanger sequencing | SE |  |
|  | (2) M Sekine et al. (2001), *Clin Cancer Res* | Sanger sequencing | SE |  |
|  | (3) K Sugano et al. (2008), *Cancer Sci* | MLPA, Sanger sequencing | SE |  |
| Korea | (1) KJ Eoh et al. (2018), *Cancer Res Treat* | NGS panel, Sanger sequencing | SE |  |
|  | (2) H Kim et al. (2012), *Breast Cancer Res Treat* | DHPLC, CSCE, CSGE, Sanger sequencing | SE |  |
|  | (3) BS Kwon et al. (2019), *Cancer Res Treat* | DHPLC, Sanger sequencing | HT, SE | Primary breast cancer; FDR/SDR with BC/OC |
|  | (4) B Park et al. (2017), *Breast Cancer Res Treat* | MLPA, Sanger sequencing | SE |  |
| Malaysia | (1) E Thirthagiri et al. (2008), *Breast Cancer Res* | MLPA, Sanger sequencing | SE |  |
|  | (2) XR Yang et al. (2017), *Breast Cancer Res Treat* | MLPA, Sanger sequencing | HT, SE | Family history of BC |
| Saudi Arabia | (1) R Bu et al. (2016), *Int J Cancer* | Sanger sequencing | HT | FDR/SDR with BC/OC |
| Spain | (1) E Beristain et al. (2007), *Breast Cancer Res Treat* | CSGE, Sanger sequencing | SE |  |
|  | (2) P Blay et al. (2013), *BMC Cancer* | MLPA, Sanger sequencing | SE |  |
|  | (3) I de Juan et al. (2015), *Fam Cancer* | CSCE, CSGE, HDX, HPLC, HRM, MLPA, NGS, Sanger sequencing | SE |  |
|  | (4) S de Sanjose et al. (2003), *Int J Cancer* | DHPLC, HDX, Sanger sequencing | SE |  |
|  | (5) O Díez et al. (2003), *Hum Mutat* | CSGE, DGGE, PTT, Sanger sequencing, SSCP | SE |  |
|  | (6) G Llort et al. (2002), *Hum Mutat* | Sanger sequencing | SE |  |
|  | (7) MD Miramar et al. (2008), *Breast Cancer Res Treat* | DHPLC, MLPA, Sanger sequencing | SE |  |
|  | (8) A Ruiz de Sabando et al. (2019), *BMC Cancer* | MLPA, Sanger sequencing | SE |  |
| UK | (1) Anglian Breast Cancer Study Group (2000), *BR J Cancer* | HDX, Sanger sequencing | SE |  |
|  | (2) VM Basham et al. (2002), *Breast Cancer Res* | HDX, Sanger sequencing, SSCP | SE |  |
|  | (3) J Peto et al. (1999), *J Natl Cancer Inst* | CSGE, Sanger sequencing | SE |  |
| USA | (1) EB Claus et al. (2005), *JAMA* | Sanger sequencing | HT, SE | FDR with BC |
|  | (2) AW Kurian et al. (2009), *Cancer Epidemiol Biomarkers Prev* | 2D Gene Scanning, Exon Grouping Analysis, Sanger sequencing | HT | BC at <35 years; bilateral BC at <50 years; prior OC or childhood cancer; FDR with BC/OC |
|  | (3) AM Martin et al. (2001), *J Clin Oncol* | CSGE, HDX, Sanger sequencing | SE |  |
|  | (4) Z Nahleh et al. (2014), *Am J Cancer Res* | BART, Sanger sequencing | SE |  |

|  |  |  |  |
| --- | --- | --- | --- |
| (5) R Nanda et al. (2005), *JAMA* | DHPLC, SSCP, Sanger sequencing | SE |  |
| (6) T Pal et al. (2005), *Cancer* | Sanger sequencing | HT, SE | FDR/SDR with BC/OC |
| (7) T Pal et al. (2015), *Cancer* | MLPA, Sanger sequencing | SE |  |
| (8) JN Weitzel et al. (2005), *Cancer Epidemiol Biomarkers Prev* | Sanger sequencing | SE |  |

†Definition of “likely hereditary” only provided for those countries where pathogenic BRCA1/2 prevalence estimates were calculated using the Horvitz-Thompson pseudo-likelihood function
BART: BRCA Analysis Rearrangement Testing, BC: breast cancer, CSCE: conformation sensitive capillary electrophoresis, CSGE: conformation sensitive gel electrophoresis, DGGE: denaturing gradient gel electrophoresis, DHPLC: denaturing high performance liquid chromatography, FDR: first-degree relative, HDX: heteroduplex analysis, HPLC: high performance liquid chromatography, HRM: high resolution melting analysis, HT: Horvitz-Thompson prevalence calculation, MBC: male breast cancer, MLPA: multiplex ligation-dependent probe amplification, NGS: next generation sequencing, OC: ovarian cancer, PTT: protein truncation test, SDR: second-degree relative, SE: species estimation, SSCP: single strand conformation polymorphism

The estimated total number of unique pathogenic variants ranged from 137 (95% CI: 90‥252) in Argentina to 1,153 in China (95% CI: 844‥1,466) and are shown in Table 2. Chao and pnpml estimates were calculated for 12 countries. The pnpml estimates were consistently equal to or greater than the Chao estimates, as expected. The median Chao estimate was 364, and study samples consisted of 13% to 58% of the total expected variants, predicting 34% of the total variants on average. The median pnpml estimate was 395, and study samples consisted of 12% to 55% of the total expected variants, predicting 31% of the total variants on average. Conversely, the average proportion of missing variants was 69%, with studies expected to be missing 45% to 88% of variants. While China had both the largest number of unique variants sampled and the highest Chao and pmpml estimates, when all countries were considered, having a larger sample of unique variants did not always result in larger estimates. The mixed Poisson distributions of variant frequency estimated via the pnpml method are seen in Fig 1 and the parameters that make up these mixtures are listed in S3 Table. These estimated distributions indicate a higher proportion of rare variants compared to recurrent variants making up the pathogenic burden.

**Table 2:**
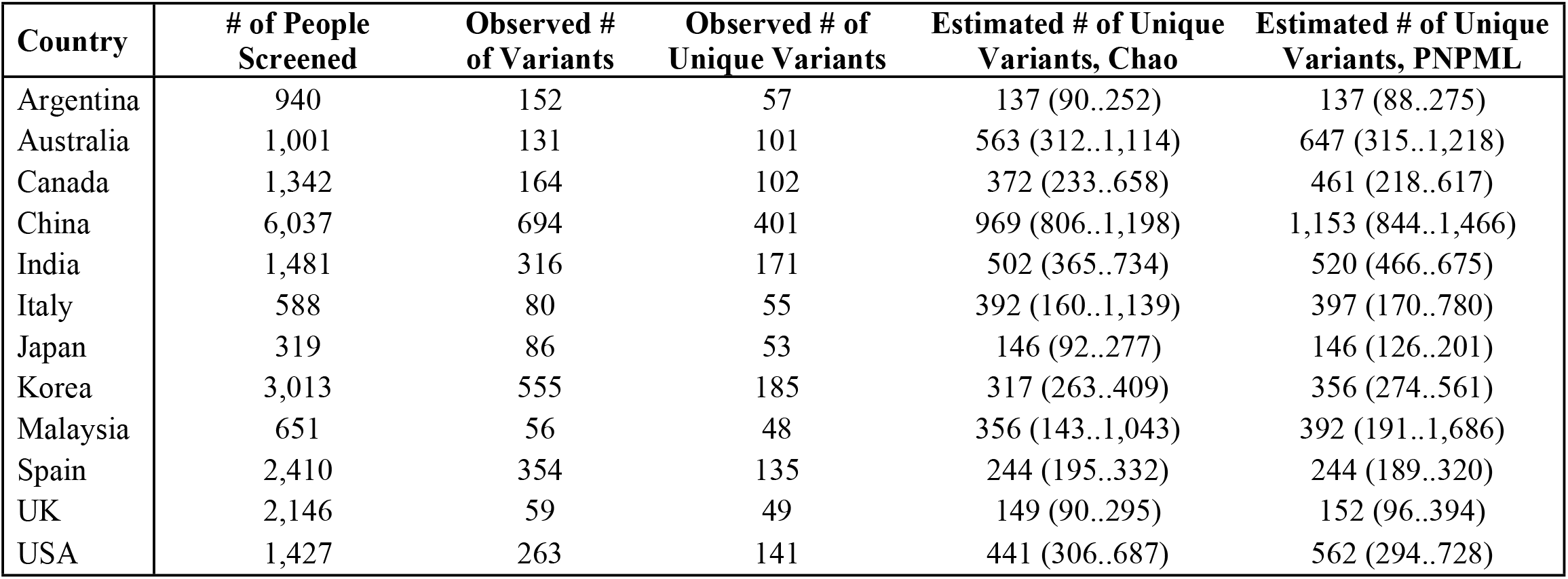
Estimates for the total number of unique pathogenic BRCA1/2 variants in different countries using Chao and PNPML estimators.

**Fig 1.**
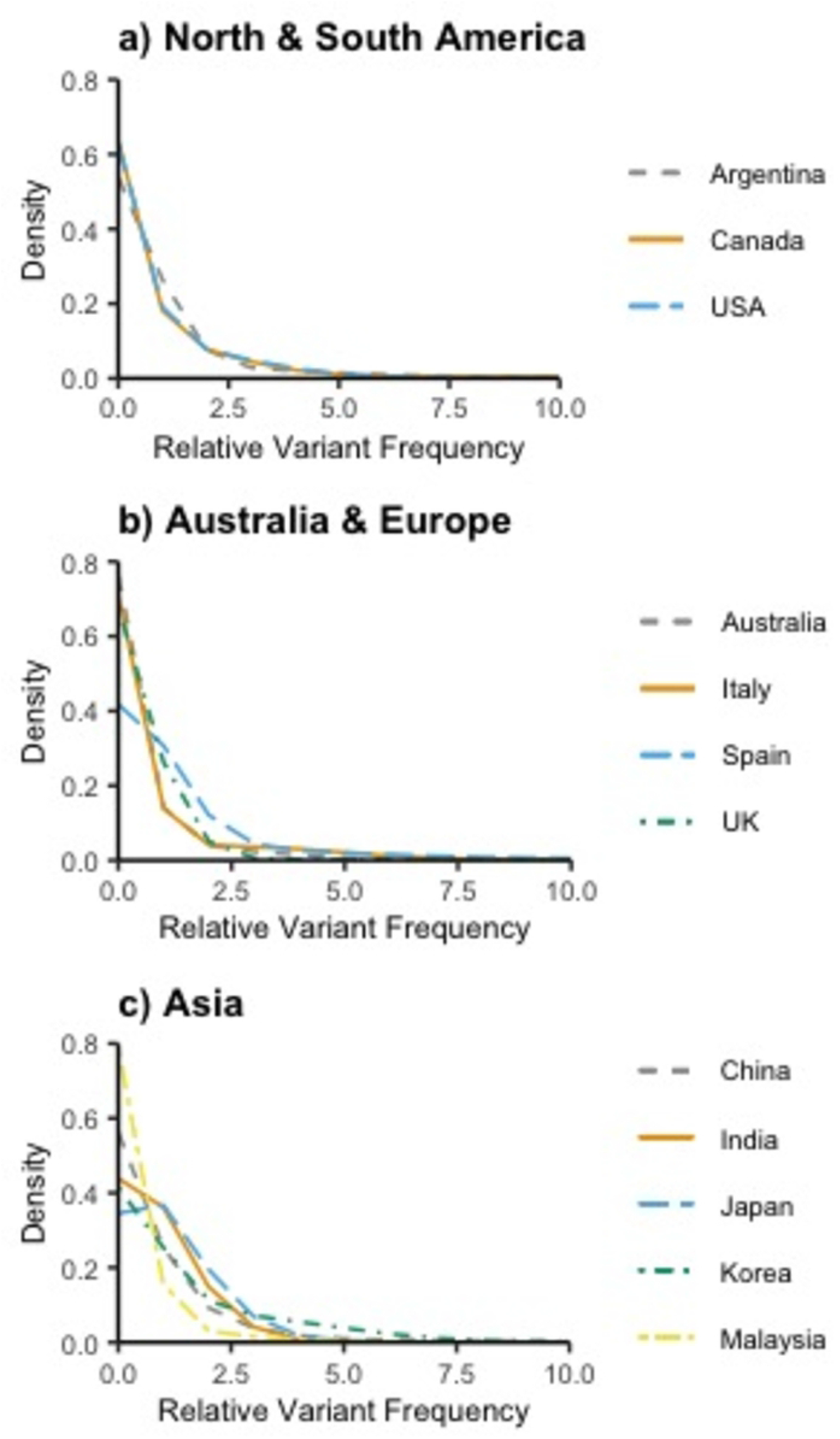
Normalized pathogenic variant frequencies in different locations globally.

Chao and pnpml estimates were calculated for 5 populations from the gnomAD data: African, American, East Asian, European, and South Asian (S4 Table). The estimated total number of unique pathogenic variants ranged from 190 (95% CI: 98‥447) for the African population to 850 (95% CI: 807‥954) for the European population. The median Chao estimate was 494. The median pnpml estimate was 558 and gnomAD data consisted of 8% to 30% of the total expected variants, predicting 23% of the total variants on average.

Estimates of the prevalence of *BRCA1/2* pathogenic variant carriers ranged from 0.09% (95% CI: 0.001, 9.42) (Denmark) to 1.05% (95% CI: 0.06, 18.58) (Austria) with a median estimate of 0.38% (IQR: 0.4) (Table 3). The median prevalence estimate was 0.64% (IQR: 0.25) among samples of breast cancer patients and 0.27% (IQR: 0.195) among samples of ovarian cancer patients. Although the prevalence point estimates were similar across countries, confidence intervals varied widely.

**Table 3:**
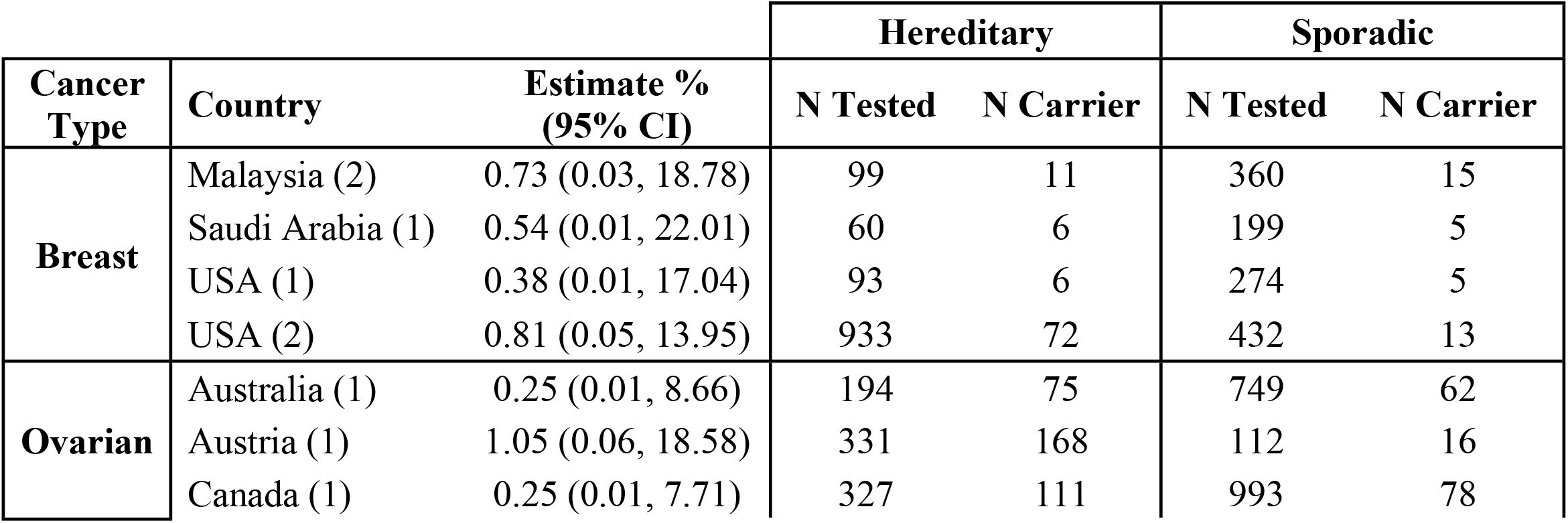
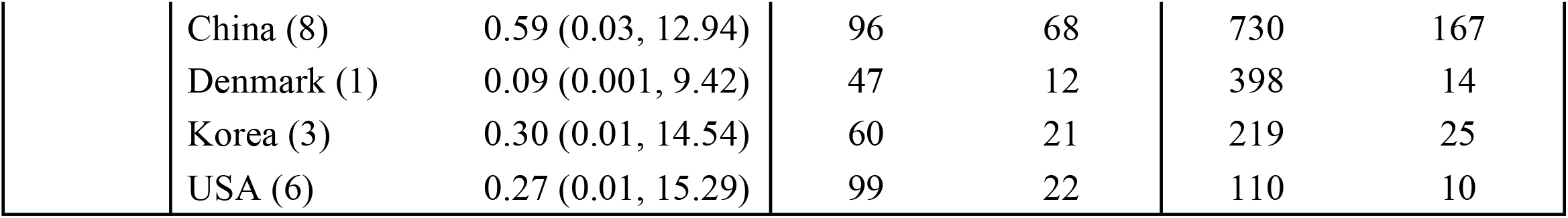
Horvitz-Thompson estimates for the prevalence of *BRCA1/2* carriers in various countries depending on cancer type.

| Cancer Type | Country | Estimate %<br>(95% CI) | Hereditary |  | Sporadic |  |
| --- | --- | --- | --- | --- | --- | --- |
|  |  |  | N Tested | N Carrier | N Tested | N Carrier |
| Breast | Malaysia (2) | 0.73 (0.03, 18.78) | 99 | 11 | 360 | 15 |
|  | Saudi Arabia (1) | 0.54 (0.01, 22.01) | 60 | 6 | 199 | 5 |
|  | USA (1) | 0.38 (0.01, 17.04) | 93 | 6 | 274 | 5 |
|  | USA (2) | 0.81 (0.05, 13.95) | 933 | 72 | 432 | 13 |
| Ovarian | Australia (1) | 0.25 (0.01, 8.66) | 194 | 75 | 749 | 62 |
|  | Austria (1) | 1.05 (0.06, 18.58) | 331 | 168 | 112 | 16 |
|  | Canada (1) | 0.25 (0.01, 7.71) | 327 | 111 | 993 | 78 |

|  |  |  |  |  |  |
| --- | --- | --- | --- | --- | --- |
| China (8) | 0.59 (0.03, 12.94) | 96 | 68 | 730 | 167 |
| Denmark (1) | 0.09 (0.001, 9.42) | 47 | 12 | 398 | 14 |
| Korea (3) | 0.30 (0.01, 14.54) | 60 | 21 | 219 | 25 |
| USA (6) | 0.27 (0.01, 15.29) | 99 | 22 | 110 | 10 |

The most recurrent pathogenic variants in the included countries are listed in Table 4. In each country, the most recurrent variant made up between 5% to 16.4% of the total observed variants and the majority of the most recurrent variants were located in *BRCA1. BRCA1* c.68_69del was seen commonly in 4 of the listed countries: Argentina, Canada, India, and the USA. Assuming that the overall prevalence of *BRCA1/2* pathogenic variant carriers is 0.62%, India likely has the highest number of people carrying this variant, with an estimated 1,233,236 (95% CI: 920,037‥1,604,558) affected, while the USA has the second highest number of carriers for this variant, with 255,353 (95% CI: 179,088‥349,425) affected. Another variant, *BRCA1* c.5266dup, was also observed commonly in 5 of the listed countries: Argentina, Australia, Canada, Italy, and the USA. The USA is estimated to have the most individuals with this variant, with 123,808 (95% CI: 71,635‥197,200) carriers.

**Table 4:** Most common pathogenic BRCA1/2 variants in different countries.

| Country | Common Variants | # of Occurrences (% of Observed) | Estimated # of Carriers (95% CI) per 100,000 people* | Estimated # of Carriers (95% CI) Overall† |
| --- | --- | --- | --- | --- |
| Argentina | *BRCA1* c.68_69del | 25 (16.4) | 102 (68, 145) | 45,826 (30,482..64,975) |
|  | *BRCA2* c.5946del | 21 (13.8) | 86 (55, 127) | 38,494 (24,408..56,672) |
|  | *BRCA1* c.5266dup | 14 (9.2) | 58 (32, 93) | 25,663 (14,294..41,710) |
|  | *BRCA1* c.211A>G | 11 (7.2) | 45 (23, 78) | 20,164 (10,226..35,051) |
| Australia | *BRCA1* c.5266dup | 8 (6.1) | 38 (17, 73) | 9,604 (4,199..18,368) |
| Canada | *BRCA1* c.5266dup | 11 (6.7) | 42 (22, 73) | 15,632 (7,924..27,221) |
|  | *BRCA1* c.68_69del | 10 (6.1) | 38 (19, 68) | 14,211 (6,899..25,473) |
| China | *BRCA1* c.5470_5477del | 38 (5.5) | 34 (25, 47) | 474,499<br>(337,968..644,738) |
| India | *BRCA1* c.68_69del | 46 (14.6) | 91 (68, 118) | 1,233,236<br>(920,037..1,604,558) |
|  | *BRCA1* c.5074+1G>A | 22 (7.0) | 44 (28, 65) | 589,809<br>(373,606..876,831) |
| Italy | *BRCA1* c.1380dup | 6 (7.5) | 47 (18, 97) | 28,039 (10,468..58,358) |
|  | *BRCA1* c.5266dup | 6 (7.5) | 47 (18, 97) | 28,039 (10,468..58,358) |
|  | *BRCA1* c.3756_3759del | 4 (5.0) | 32 (9, 77) | 18,693 (5,160..46,021) |
|  | *BRCA2* c.6468_6469del | 4 (5.0) | 32 (9, 77) | 18,693 (5,160..46,021) |
| Japan | *BRCA1* c.188T>A | 13 (15.1) | 94 (52, 152) | 118,337 (64,976..191,484) |
|  | *BRCA1* c.2800C>T | 11 (12.8) | 80 (41, 135) | 100,132 (51,355..170,112) |
| Korea | *BRCA2* c.7480C>T | 51 (9.2) | 57 (43, 74) | 29,461 (22,186..38,151) |
| Malaysia | *BRCA2* c.8961_8964del | 4 (7.1) | 45 (13, 108) | 14,150 (3,923..34,250) |
|  | *BRCA2* c.5353_5354del | 3 (5.4) | 34 (7, 93) | 10,612 (2,219..29,456) |
| Spain | *BRCA2* c.2808_2811del | 32 (9.0) | 57 (39, 78) | 26,385 (18,301..36,543) |
|  | *BRCA2* c.9026_9030del | 23 (6.5) | 41 (26, 60) | 18,964 (12,143..27,991) |
|  | *BRCA2* c.211A>G | 18 (5.1) | 32 (19, 50) | 14,842 (8,874..23,117) |
| UK | *BRCA1* c.4065_4068del | 3 (5.1) | 32 (7, 88) | 21,070 (4,393..58,634) |
| USA | *BRCA1* c.68_69del | 33 (12.5) | 78 (55, 107) | 255,353<br>(179,088..349,425) |
|  | *BRCA1* c.5266dup | 16 (6.1) | 38 (22, 61) | 123,808 (71,635..197,200) |
|  | *BRCA2* c.5946del | 15 (5.7) | 36 (21, 58) | 116,070 (65,734..187,839) |

†Using prevalence estimate of 0.62

## Discussion

Species richness methods from ecology can provide informative estimates of “variant richness” or the number of missing pathogenic variants in a location and the relative frequency of these variants. Results from the unique variant estimation indicate that for the included countries, between 45% and 88% of pathogenic *BRCA1/2* variants have yet to be observed in research studies. While different countries have different variant frequencies, all countries appear to have many more rare pathogenic variants compared to recurrent variants. This suggests that most of the variants that have not yet been identified in each studied country will be rare.

Species richness methods applied to gnomAD data additionally suggest that many pathogenic *BRCA1/2* variants are still missing in research data. The prediction of fewer African *BRCA1/2* variants may be due, in part, to smaller sample size, but may also be a reflection of more prominent recent population growth in other populations as dated pathogenic founder variants occurred relatively recently [24,25,26,27]. While gnomAD is broken out by ancestry rather than nationality, from a public health standpoint, nationality may be a more straightforward and useful metric around which to design screening strategies.

The *BRCA1/2* prevalence point estimates reported here are similar to those previously reported by population genetic screening studies [2,3,4]. We observed a median prevalence estimate of 0.64% and 0.27% using samples of breast and ovarian cancer patients, respectively. In comparison, the Healthy Nevada Project reported a 0.66% prevalence for pathogenic *BRCA1/2* variant carriers [3] the Geisinger MyCode Community Health Initiative reported 0.5% [4] and the Bio*Me* Biobank reported 0.7% [2]. Our results have greater uncertainty and wider confidence intervals due to smaller samples in the international set of studies we included compared to other studies that present prevalence estimates. For a small number of included studies, cases with unknown heredity were excluded, while cases with complete heredity information were used for estimation. This may have resulted in sampling bias [28,29,30] perhaps making estimates for these countries appear larger because cases with strong family history are less likely to have unknown hereditary information compared to sporadic cases. In addition, different definitions for hereditary and sporadic cancers across studies may have impacted results. However, similar overall results between our study and others suggest that the prevalence of *BRCA1/2* pathogenic variants is relatively consistent despite there being unique recurrent and rare variants represented in different populations. This observation is consistent with an assumption of similar mutation rates in different populations and with documented global population growth [31].

The largest number of estimated pathogenic *BRCA1/2* carriers are seen in countries with large populations, such as China and India. *BRCA1* c.68_69del is a commonly observed variant in several countries and may be the most recurrent *BRCA1/2* variant globally. Although it is popularly known because of its high frequency in individuals of Ashkenazi Jewish descent [32], the reason for its high estimated occurrence globally is primarily because of its apparent frequency in India. Individuals with this variant may mistakenly believe they have Jewish ancestry, even though this variant has been shown to occur on a different haplotype [33]. Despite being recognized as a highly observed variant in India [34,35,36,37], the risk implications of *BRCA1* c.68_69del for individuals of Asian Indian ancestry are not currently acted upon clinically. The list of *BRCA1/*2 variants expected to be most recurrent in different parts of the world (Table 4) highlights situations like this and suggests that there may still be recurrent variants in less well studied countries with significant and unrecognized clinical implications.

Our study does not represent all literature on *BRCA1/2* and includes a limited number of countries because species richness methods cannot be accurately applied for countries where only a small number of pathogenic *BRCA1/2* variants have been observed [11]. Therefore, we limited estimations about missingness and the frequency of missing variants to only those locations where a modest amount of research has already been conducted. Even for countries included, the samples used may not be representative of the country in its entirety [7,38,39,40] and different sequencing strategies, enrollment criteria, and sampling strategies across studies may have biased the results for some countries. Furthermore, differences in the number of expected variants between populations may be attributable to estimation error or size of population, rather than biological differences in underlying population genetics. These limitations of the current literature are consistent with and strengthen our conclusion that there is a large amount of missing information about *BRCA1/2* pathogenic variation globally.

Despite being two of the most studied genes in the world, much information is still missing about pathogenic variation in *BRCA1/2*. Multiple studies examining *BRCA1/2* variation that we observed but did not include in our study only sequenced a small set of variants due to cost constraints [41,42]. While such population-based strategies targeting specific variants commonly observed among cases have been proposed as potentially being more cost-efficient [5,6], these strategies assume that the variants observed in a small subset of individuals will represent a large portion of variants observed throughout the population. The species richness methods presented here provide a more rigorous statistical means to evaluate if targeted assays will achieve the desired sensitivity in a given population. We suggest that future surveys of genetic variation also model variant richness as we have described. This will provide a picture of what is observed with estimates of what is still unknown.

## Data Availability

Data generated or analyzed during this study are included in this published article and its supporting information files. Additional datasets used for analysis are available in The Genome Aggregation Database (gnomAD), https://gnomad.broadinstitute.org/downloads.

## Acknowledgements

None.

## Supporting information

**S1 Fig. Flow diagram of study search and selection procedure**.

**S1 Table. Data used in variant estimation calculations**.

**S2 Table. Data used in prevalence calculations**.

**S3 Table. Parameters for compound Poisson distributions fit for the PNPML estimates using country data**.

**S4 Table. Estimates for the total number of unique pathogenic BRCA1/2 variants in different gnomAD populations using Chao and PNPML estimators**.

